# Covid-19 Per Capita Fatality Rate: A Path Analysis Model

**DOI:** 10.1101/2021.09.22.21263976

**Authors:** Michael Penuliar, Candice Clark, Debra Curti, Miguel Carrasco, Catherine Hudson, Billy Philips

## Abstract

**Objectives:** Various individual factors have been shown to influence Covid-19 mortalities, but these factors do not exist in isolation. Unique to this study is a multivariate approach that has yet to be fully explored by previous research. Using an interconnected multifactor model, this work investigated social determinant, geographic, prior health, and political behavioral factors likely to influence Covid-19 per capita fatalities in Texas.

**Methods:** County-level income, rurality, insurance, health status, 2020 presidential vote percentage, and fatality rate data were collected and analyzed in a path analysis model with Covid-19 per capita fatalities as the key variable of interest.

**Results:** The analysis found strong support for the proposed model structure (*R*^2^ = 37.6%). The strongest overall effects on the Covid-19 per capita fatality rate came from income levels and voting behaviors.

**Conclusion:** The model explained a substantial amount of variability in mortalities attributed to Covid-19. Socioeconomic and political factors provided the strongest contribution to the per-capita Covid-19 death rate, controlling for the other variables studied. The Covid-19 pandemic was highly politicized by various leaders and media outlets. The current analysis showed that political trends were one of the key overall factors related to Covid-19 mortality. The strongest overall factor was median income. Income is used to enhance one’s current health or acquire adequate treatment which may safeguard people from the most severe effects of Covid-19. Counties with lower income levels had higher rates of Covid-19 per capita fatalities.

## Introduction

Major causative reasons for the Covid-19 case and fatality rates have been proposed by many scholars. For example, income, geographic location, insurance rate, health status, and politicization are some factors suggested as individual contributors that spur both case and fatality rates, but no study has looked at the effect of these types of variables in unison within a multifactor model. This study embarked on the notion that these factors reside in an interconnected framework and produce both direct and indirect effects on our key variable of interest, the Covid-19 fatality rate.

First, regarding income level and its relationship to Covid-19 fatalities, a study investigating Covid-19 hospitalization factors found that those who resided in higher income zip codes, compared to lower income zip codes, were less likely to be hospitalized as a result of the virus,^1,2^ signifying that the benefits afforded by a higher income act as a protective buffer to the most severe effects of the Covid-19 virus. Other researchers have also found a connection between median household income and increased Covid-19 fatalities,^3^ and they also specify that access to healthcare is scarcer in lower income areas which may lead to higher fatality rates. A lack of access to care is especially true for rural areas another factor in our proposed framework.

Rural counties are far more likely to contain individuals who are more advanced in age, have less access to care and lower median incomes, and are more likely to have pre-existing conditions.^4^ These elements can contribute to an increased risk of Covid-19 fatalities or severe cases. Regarding good public health practices, researchers have also found that rural residents were less likely to have worn a mask, practiced good sanitation habits, avoided dining out, or worked from home during the pandemic.^5^ Together, these behaviors have the potential to increase the exposure of the disease and thus a community’s fatality rate as well.

A third factor to consider is the lack of health insurance. In the United States, a lack of health insurance is associated with higher mortality rates.^6^ Regarding income, an aforementioned factor in our proposed framework, researchers have found an association between lower median family incomes and an increased amount of people without health insurance coverage,^7,8^ giving further credence to an interconnected modeling approach. A lack of health insurance may also result in a reduced ability or desire to obtain adequate care, preventative or otherwise, leading to overall poor health, yet another factor in our multi-variable Covid-19 fatality framework.

Poor initial health is often reasoned as a major catalyst for a Covid-19 fatality. Concerning health and insurance, one study found a lack of insurance was not only connected to increased all-cause and numerous other mortality types, but also related to inflammation inducing lifestyle factors such as poor diet, lack of exercise, obesity, and smoking and alcohol use,^9^ which may contribute to a higher fatality risk in Covid-19 patients. Similarly, comorbid heart and lung conditions such as hypertension, cardiovascular disease, or chronic obstructive pulmonary disease are seen as major initial risk factors for increased fatality rates and more severe prognoses of Covid-19 patients.^10^

A final factor that this research considered was the politicization of the pandemic and the Covid-19 virus. Science communication research found that politicians were featured more heavily than scientists in articles on traditional news outlets like the *USA Today* and the *Washington Post*,^11^ which likely biased some of the public’s information regarding the Covid-19 health crises and divided public opinion. Researchers have also posited that politicization from political leaders and the media influenced community risk perceptions. Specifically, one study found that as the proportion of votes for Trump from the 2016 United States presidential election increased there were decreased internet searches for Covid-19 related information and minimal change in visitation rates to businesses relative to before the pandemic.^12^ Research has also found a relationship between conservative media consumption, reductions in physical distancing, and increased case and fatality rates of a given area.^13^

These factors discussed do not exist in isolation, and they likely influence each other and Covid-19 fatalities as a whole. This research sought to examine the aggregate effect of the aforementioned socially determinant, behavioral, and geographic factors and their relationship to the Covid-19 fatality rate in Texas, a large and diverse state that is the second most populous and second largest state by land area within the United States. To date and to our knowledge, no significant research has been conducted in creating a multifactor path model to investigate the major root causes of the Covid-19 fatality rate.

It was hypothesized that the median household income, uninsured rate, residents’ health status, rurality, and voting trends would collectively predict Covid-19 fatality rates. All 254 counties in Texas were aggregated and analyzed via path analysis which describes the direct, indirect, and overall dependencies among a set of variables and the strength of those relationships. Of the given variables, this study posited that the strongest overall effects on Covid-19 fatality rates would derive from median household income and voting behavior considering the strength of politicization during the Covid-19 pandemic and that income is used to finance adequate personal and preventative health in order to stymie the most severe effects from the disease.

## Methods

The data collected for this study was readily available online for public use and thus an Institutional Review Board evaluation was not needed. As this study collated and analyzed public data, there were no human subjects and thus no informed consent was necessary. Data was analyzed using the path analysis software, SPSS Amos (v 26). The Robert W. Johnson’s County Health Rankings’ 2020 dataset provided county-level data for the State of Texas on median household income (Median Income), uninsured rate (Uninsured), and the percentage of adults considered to be in poor or fair health (Poor/Fair Health).^14^ Rural-Urban Continuum Code (RUCC) data was collected from the United States Department of Agriculture, counties were rated on a scale of 1-9, with 1 as the upper limit of urbanization, i.e., highly metropolitan, and 9 as the lower limit of urbanization, i.e., highly rural.^15^ *The New York Times* provided the county-level Covid-19 fatality data which was retrieved on February 23, 2021.^16^ Covid-19 fatality data was adjusted to a per 100,000 rate (Covid-19 Fatalities per Capita). Finally, Texas county 2020 presidential voting data was collected from NBC News which provided the percent of a county’s population that voted for the Democratic candidate, Joe Biden, during that particular election cycle (Percent 2020 Democrat Vote).^17^ This variable was chosen as the percentage of the vote received for the 2020 presidential republican candidate, Donald Trump (Percent 2020 Republican Vote) was approximately equal to 1 - Percent 2020 Democrat Vote across all Texas counties, and thus these two variables were nearly interchangeable. Upon observing the data collated for this study, there was no missing data to report for the *n* = 254 Texas counties.

The analytical technique used for this study was path analysis, often considered a specialized form of multiple regression or structural equation modeling. This type of analysis allows researchers to chart and analyze potential direct and indirect relationships amongst a multitude of variables based on hypothesized causal structures informed by theory or previous research. The five independent variables in our analysis were Uninsured, Median Income, Poor/Fair Health, RUCC, and Percent 2020 Democrat Vote. The dependent variable was Covid-19 Fatalities per Capita. These variables were inserted into our hypothesized path analysis structure and analyzed.

For our path structure, it was hypothesized that Covid-19 Fatalities per Capita, would be directly predicted by Median Income, Poor/Fair Health, RUCC, and Percent 2020 Democrat Vote. It was also hypothesized that Poor/Fair Health and Percent 2020 Democrat Vote would be directly predicted by RUCC, Uninsured, and Median Income. It was believed that Uninsured would be predicted by RUCC and Median Income. Finally, when considering the total effects, we predicted that Median Income and Percent 2020 Democrat Vote would have the strongest total effects on Covid-19 Fatalities per Capita.

## Results

An exploratory factor analysis and a Harman’s one-factor test found that all variables loaded onto more than one factor and constraining the analysis to one factor resulted in a variance explained of less than 50%, and thus common method variance was not an issue.^18,19^ Multicollinearity was not an issue as VIF < 5, Tolerance >.2, and there were no correlations above *r* = |.90| between all bivariate correlation variable combinations.^20,21,22^ Finally, there were no normality issues as all variable skews were less than 2 and all coefficients of kurtosis were less than 7.^23^

Path model structure, standardized direct path estimates, and all *R*^2^ values are shown in the Figure 1 path diagram. Table 1 details all total, direct, and indirect standardized and unstandardized effects on Covid-19 Fatalities per Capita. The fit indices used were the Root Mean Square Error of Approximation (RMSEA), Comparative Fit Index (CFI), and the Tucker-Lewis Index (TLI). Good model fit is indicated by *X*^*2*^ < 2, *RMSEA* < .06, *CFI* > .95, and *TLI* > .95. ^20,24,25^

**Table 1.** Total, Direct, and Indirect Standardized and Unstandardized Effects on Covid-19 Fatalities per Capita in Texas.

| **Effect Type on Dependent Variable** | **Median**<br>**Income** | **RUCC** | **Uninsured** | **Poor / Fair**<br>**Health** | **Percent 2020**<br>**Dem Vote** |
| --- | --- | --- | --- | --- | --- |
| Total Effect on Covid-19 Fatalities per Capita | -.413 (-3.812) | .226 (9.737) | .116 (322.523) | .180 (411.794) | -.319 (-254.775) |
| Direct Effect on Covid-19 Fatalities per Capita | -.237 (-2.184) | .115 (4.964) | .000 (.000) | .424 (971.706) | -.319 (-254.775) |
| Indirect Effect on Covid-19 Fatalities per Capita | -.176 (-1.628) | .111 (4.774) | .116 (322.523) | -.244 (-559.913) | .000 (.000) |

Note: Unstandardized estimates are in parentheses.

**Figure 1.**
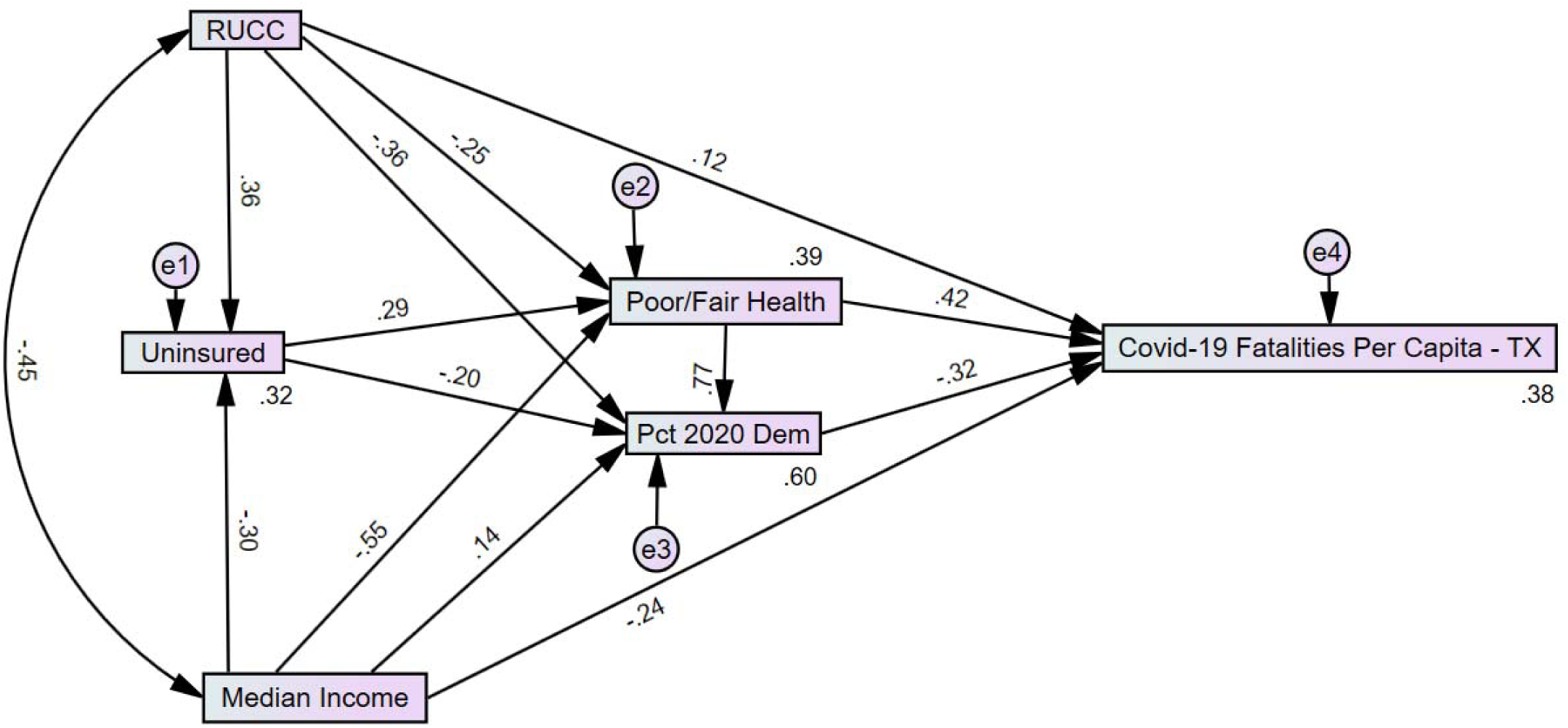
Covid-19 Per Capita Fatalities Model: Total and Indirect Standardized Effects in Texas.

The analyzed path model showed strong fit, *X*^*2*^ (1) = .33, *p* = .57, *RMSEA* < .001, *CFI* = 1.00, *TLI* = 1.02, and it predicted 37.6% of the variance in Covid-19 per Capita Fatalities in Texas. For all direct path effects, p-values were below .001 except for the effect of Median Income on Percent 2020 Democrat Vote path (*p* = .013) and the effect of RUCC on Covid-19 Fatalities per Capita path (*p* = .078) which was marginally significant.

Upon observing the model’s direct effects on the Covid-19 fatality rate, Median Income and Percent 2020 Democrat Vote were negatively related to Covid-19 Fatalities per Capita. As the median income and the percent of the vote allotted to the Democratic candidate of the 2020 presidential election in a county decreased, Covid-19 Fatalities per Capita increased. In comparison, as RUCC and Poor/Fair Health increased within a given county so did Covid-19 Fatalities per Capita. Of all the direct pathways to Covid-19 Fatalities per Capita, Poor/Fair health showed the strongest direct relationship followed by Percent 2020 Democrat Vote.

All indirect and total path estimates were significant at *p* < .01. Median Income and Poor/Fair Health both had negative indirect effects on Covid-19 Fatalities per Capita. In comparison, RUCC and Uninsured had positive indirect effects on Covid-19 Fatalities per Capita. The strongest indirect paths were Poor/Fair Health followed by Median Income. After accounting for both direct and indirect effects, the factors that had the strongest total effects on Covid-19 Fatalities per Capita were Median Income followed by Percent 2020 Democrat Vote, which satisfied our prediction.

## Discussion

The current study provided a less myopic view of Covid-19 mortality by examining a multitude of variables. The five-predictor path model explained a considerable amount of variability in the State of Texas’ Covid-19 per capita fatality rate. It also replicates and extends many of the standalone findings of previously mentioned research such as the relationship between the uninsured rate or median income on health and mortality.^6,7,8^

In the current study, when observing the strongest direct effects, counties worse in overall health and those with decreased Democratic voting percentages had higher Covid-19 fatalities relative to their population. Regarding total effects, which considers all prior mediating effects, a county’s median income followed by their Democratic voting percentages were the strongest Covid-19 fatality rate predictors. This finding occurred because the Percent 2020 Democrat Vote mediation inversed and weakened the total Poor/Fair Health effect.

Within the path analysis, there was a strong positive relationship between Poor/Fair Health and Percent 2020 Democrat Vote. This relationship was also the strongest in the entire path model structure. Counties with an increased amount of people in poor or fair health were more likely to vote for the Democratic candidate, presumably because of a general perception that the United States’ Democratic platform, relative to other party options, is more likely to expand or create social safety nets such as single-payer health insurance.^26^ Importantly, counties that were overall more likely to vote Democrat, taking into account all proceeding variables, were less likely to have increased Covid-19 fatality rates. The effect of these relationships thus reduced the total effect of the Poor/Fair Health factor on the Covid-19 Fatalities per Capita rate. One reason is that areas that are heavily Democrat tend to center or surround more urban clusters where access to care is more readily available, and the opposite is true for rural clusters.^27^

Concerning the strongest overall factor in the model, Median Income, there is substantial previous research on the relevancy of income on the role of one’s health. Research has shown that income, especially those in the lowest order of the income distribution, and income inequality overall have a strong relationship with increased overall mortality.^28,29^ This has also been seen in other health conditions. For instance, previous patient studies have found that lower-income stroke and cystic fibrosis patients had increased mortality rates as well.^30,31^ In the United States, income is a major factor in health insurance status as it is directly tied to most people’s employment and level of employment. Health insurance status then drives one’s personal health status which ultimately effects mortality risk.

In the examinations of both total and direct effects, voting behavior was the second strongest path analysis factor regarding the Covid-19 per capita fatality rate. To date, this is the first known study of its kind to directly examine the relationship between voting behavior from the 2020 United States presidential election and Covid-19 fatalities as well as their relationship in a larger structure. Related research on 2016 presidential voting data found a negative relationship between Covid-19 case rates and the support for the Democratic candidate and the opposite was true for the Republican candidate,^32^ which lends further credence that the outcome of the pandemic is at least partially due to politicization. Related research has shown that political conservative attitudes to be associated with decreased adherence to public health guidelines as well as beliefs of decreased vulnerability to Covid-19, decreased perception of Covid-19 severity, and an endorsement that the virus’ effects have been exaggerated.^33,34^

## Limitations and Future Directions

For future work, a comparative fatality rate model of a similar structure to assess other infectious diseases such as the flu may help further gauge the model’s accuracy and generalizability. Conceivably a less politicized public health matter such as the flu may have a substantially weaker relationship with voting behaviors. Though the current work focused on the State of Texas and found strong results, researchers should replicate this model with other states or the United States as a whole to see what factors may be the most relevant to other geographic areas or if there are other factors yet to be considered. This work was also concluded prior to the wide scale distribution of Covid-19 vaccinations in the United States and the Delta variant surge. Future researchers may assess if vaccine distribution and variant surges alter or strengthen some of the associations made in this analysis. Finally, experimental or quasi-experimental studies may better determine causality based on our or future path analysis chains related to Covid-19 fatalities.

## Conclusion

The Covid-19 per Capita Fatality rate is derived from a unique and complex structure. As expected, factors related to one’s health and income were tied to the fatality rate, but unique to this structure is the strong relationship between Covid-19 fatalities and individual political behaviors. This politicization potentially contributed to unnecessary excess mortality, and it is imperative to safeguard against politicization for all current and future public health crises.

## Data Availability

All data used for this study is publicly available online and can be analyzed by anyone using the methods detailed in this paper.

https://github.com/nytimes/covid-19-data

https://www.nbcnews.com/politics/2020-elections/texas-president-results

https://www.countyhealthrankings.org/explore-health-rankings/rankings-data-documentation

https://www.ers.usda.gov/data-products/rural-urban-continuum-codes

